# Management and outcomes of Chronic Subdural Hematoma in Africa: a protocol for a scoping review

**DOI:** 10.1101/2024.02.17.24302660

**Authors:** Victor Meza Kyaruzi, Berjo Takoutsing, Astel Dongmo, Michael Nana Yaw Yankey, Yvanah Owoundi Mbozo’o, Nathan Ezie Kengo, Fodop Samuel, Mingo Naomi Younwi, Evert Foyenka, Tcheutchoua Kathy Foka, Yab Parfait Motah, Opara Oluwamayowa, Roland Nchufor, Yannick Onana, Ngaroua, Atangana Ernestine Renée Bikono, Ignatius Esene

## Abstract

**Introduction:** Chronic subdural hematoma (cSDH) is a common neurosurgical condition characterised by the accumulation of blood between the dura mater and the arachnoid membrane. It has a significant prevalence and high mortality. Over the years, the management of cSDH has been extensively studied in high-income countries. On the other hand, Africa, which is faced with a significant neurosurgical care deficit, lacks aggregate data on the management strategies and outcomes on cSDH on this continent. This protocol aims to guide the scoping review of published studies investigating the characteristics, management strategies, and outcomes of cSDH in Africa.

**Methods and Analysis:** The scoping review protocol for the proposed study is in accordance with the Arksey and O’Malley’s framework. The objectives, eligibility criteria and search strategy were developed based on the Population, Intervention, Comparator, Outcome framework. A comprehensive search will be conducted in electronic databases, including MEDLine, African Index Medicus, PubMed, Embase, African Journals Online, and Scopus to identify relevant studies. Only peer-reviewed publications with primary data reporting on the management of cSDH in the African human population will be included. Extracted data from included articles will be presented as descriptive statistics, pooled statistics, and a narrative description where deemed necessary.

**Ethics and dissemination:** Given the non-direct involvement of human individuals, ethical approval will not be necessary. Dissemination strategies will include publication in a peer-reviewed journal, oral and poster presentations at conferences and promotion over social media.

**ARTICLE SUMMARY:** *Study strengths and limitations:*

- To our knowledge this is the first scoping review focusing on the management and outcomes of chronic subdural haematoma on the African continent.
- This protocol aims at ensuring transparency in the research methods used for the proposed review, and this will therefore reduce the chances of reviewing bias.
- The search strategy will be run in six electronic databases commonly used as repositories by journals publishing research from Africa.
- There will be no restrictions on language, and publication date during the screening process.
- Unpublished studies will not be sought.
- There will be no formal assessment on the quality of the included studies.

## INTRODUCTION

Chronic subdural hematoma (cSDH) is a common neurological condition characterised by the accumulation of blood between the brain’s surface and its protective covering, the dura mater (1). Globally, it has an incidence between 1.72 to 20.6 per 100,000 persons per year, which increases with age (2, 3). According to the literature, many predisposing factors exist. These include more commonly advancing age (≥ 65 years), and the use of antiplatelet or anticoagulant medications (3). Less commonly, chronic alcoholism, epilepsy, and coagulopathy (4). In west Africa and other parts of the world, there is a relatively higher preponderance of males than females in cSDH (5). This may be attributed to a greater exposure of males to injury.

Despite how common and the wide prevalence of cSDH, there remains a lack of consensus regarding numerous aspects of its clinical management (6). In Africa in particular, there have been recent reports of changing trends of this condition (7). Also, despite the advances in neurosurgical diagnostic capacity, Africa faces unique challenges in the management of neurosurgical conditions, including limited resources, inadequate access to neurosurgical services, and healthcare disparities (8).

## STUDY AIMS

This study describes the protocol for conducting scoping review aiming at exploring the management strategies and outcomes of cSDH in Africa. The information gained will help to identify key priorities and the establishment of larger scale research with greater evidence.

### Primary aim

1. Describe the various treatment options for cSDH on the African continent.

### Secondary aim

2. Assess the methods of diagnosis of cSDH in Africa.
3. Evaluate the clinical outcomes primarily defined as, mortality, recurrence and complication rates, functional outcome, Length of hospital stay, and clinical state on discharge for patients suffering of cSDH in Africa.
4. Describe the demographic and etiologic characteristics of cSDH in Africa.

## METHODS

The planned review will be developed following the five stages of the Arksey and O’Malley’s framework for scoping review studies (9). This protocol is designed in accordance with the Preferred Reporting Items for Systematic Reviews and Meta-Analyses (PRISMA) of Protocols (10).

### Stage 1: Determine the research question

The Patient, Intervention, Comparison, Outcome (PICO) framework was used in determining the title, objectives, and eligibility criteria of the proposed review. Our key research question is: How are cSDH managed in Africa?

### Stage 2: Identifying relevant studies

The authors defined the keywords to be used in developing the search strategy, and defining the study eligibility criteria using the PICO framework.

### Databases and search strategy

The search strategy will be developed using a combination of relevant keywords and MeSH terms related to chronic subdural hematoma, and Africa. The search strategy will be customised for each database, incorporating Boolean operators, truncation, and proximity operators as required. The following search terms will be used: (((chronic) AND (“subdural space”[MeSH] OR (subdural) AND (hemorrhag* OR haemorrhag* OR hematoma* OR haematoma* OR bleed*)) OR “hematoma, subdural, chronic”[MeSH])

AND

Africa* OR Algeria* OR Angola* OR Benin OR Botswana* OR Burkina OR Burundi* OR “Cabo Verde” OR “Cape Verde” OR Cameroon* OR “Central African Republic” OR Chad OR Comoros OR Congo* OR “Cote d’Ivoire” OR Djibouti OR Egypt* OR Guinea* OR Eritrea* OR Eswatini OR Swaziland OR Ethiopia* OR Gabon OR Gambia* OR Ghana* OR Kenya* OR Lesotho OR Liberia* OR Libya* OR Madagascar OR Malawi* OR Mali OR Mauritania* OR Mauritius OR Morocc* OR Mozambique OR Namibia* OR Niger OR Nigeria* OR Rwanda* OR “Sao Tome and Principe” OR Senegal* OR Seychelles OR “Sierra Leone” OR Somalia OR Sudan* OR Tanzania* OR Togo OR Tunisia* OR Uganda* OR Zambia* OR Zimbabwe* OR Southafrica* OR Maghreb*

NOT

(“Animals”[MeSH] NOT “Humans”[MeSH])

The literature search will be conducted without restrictions on language or date of publication in the following electronic databases: MEDLine via Ovid, African Index Medicus, PubMed, Embase, African Journals Online, and Scopus. Additional resources such as Google scholar, and references from included studies will be explored to verify that all relevant material in the literature is included.

### Eligibility criteria

The eligibility criteria was developed, and agreed by all authors following discussion and this will be used to guide the screening phase. The eligibility criteria adhere to the PICO strategy and are listed below:

#### Participants

Studies of individuals diagnosed of cSDH in Africa, irrespective of age or gender will be considered eligible for inclusion. Patients with the diagnosis of intracranial haemorrhage which is non-chronic nor located in the subdural space without an associated cSDH will be excluded. Also, studies with no disaggregate data for one particular country will be excluded.

#### Intervention/Exposure

Studies reporting on any management strategy for cSDH, including surgical interventions (e.g., burr-hole drainage, twist-drill craniostomy, craniotomy, decompressive craniectomy), and conservative management approaches (e.g., observation, medical treatment) will be included. Patients diagnosed with a cSDH whose method of diagnosis or treatment is unclear will be excluded.

#### Outcome Measures

Studies reporting on the following outcomes of cSDH will be included: (1) mortality rate (2) recurrence rate (3) complication rate (4) functional outcome (5) length of hospital stay (6) symptoms resolved/reduced/ unchanged/worsened.

##### Comparison

We shall compare the outcomes following different modalities of treatment.

#### Study Design

Only peer-reviewed publications will be included. Trials, prospective or retrospective cohort studies, case-control studies, case reports, case series, cross-sectional studies, and audits will be included. There will be no language restriction and articles will be translated where needed using online translation platforms such as DeepL Translator (https://www.deepl.com/en/translator). The following articles will be excluded; articles with no disaggregated data on cSDH, non-peer-reviewed papers/pre-prints, opinion pieces, perspectives, editorials, letters, comments, reviews, meta-analyses and guidelines.

### Stage 3: Study selection

A calibration exercise will be carried out before title and abstract screening in order to ensure adequate understanding of the eligibility criteria by study screeners. The data records identified from the search will first be downloaded from the respective databases into EndNote X9 (11). They will then be imported into COVIDENCE Systematic Review Software (http://www.covidence.org/) where deduplication, title, and abstract screening as well as full-text screening will take place (12). Each study will then be screened using title and abstract by two independent reviewers. Studies included after the first round of screening will be further screened for full-text review. Following unblinding of decisions, disagreements will be discussed amongst the reviewers and in the case of no resolution an appeal will be made to a third reviewer or the senior author (IE).

### Stage 4: Charting the data

Data from included full-text articles will be extracted into a previously made data extraction proforma on Microsoft Excel (Microsoft, Richmond, Virginia, USA). This stage will be preceded by a pilot phase where all extractors will extract data from the same ten randomly selected included articles. This phase is important to ensure reliability of the data extraction sheet, accuracy, and homogeneity in the extraction of the data. Following this phase, necessary changes will be made to the proforma before initiation of data extraction proper. Data extracted will include (i) study characteristics, (ii) patient demographics, (iii) aetiology of cSDH (iv) clinical presentation, (v) method of diagnosis (vi) treatment strategies, (vii) treatment adjuncts (viii) mortality and morbidity rates, (ix) recurrence rate, (x) functional outcome, (xi)Length of hospital admission (xii) Symptoms resolved/ reduced/ unchanged/ worsened. All extracted data will be recorded and stored in the final extraction proforma sheet and curated before formal analysis.

### Stage 5: Collating, summarising and reporting the results

Following curation of the extracted data, it will be transferred to SPSS V.26 for analysis. Pooled statistics will be calculated and presented using measures of central tendency and spread. Categorical variables will be presented as numbers and their respective proportions. Unless otherwise stated, the statistical significance will be set at p<0.05. Results will be primarily reported in tabular forms or figures, and where necessary, a narrative description of the results will be presented by grouping the data into meaningful summaries.

### Risk of Bias Assessment

Given the nature of our study and the limited evidence on the topic which we are trying to bridge, the proposed scoping review seeks to offer a comprehensive overview of the management and outcomes of cSDH in Africa. As a result, a formal risk of bias assessment is judged unnecessary.

### Ethics and Dissemination

This study will exclusively involve secondary data collection and no human participants will be involved, hence ethical approval was not required. The results from this study will be disseminated through a peer-reviewed journal. Oral and poster presentations at local, regional, national and international conferences and promotion over social media will also be done.

### Patient and public involvement

None declared

## ACKNOWLEDGEMENTS

None declared

## TWITTER

@BerjoDongmo, @ignatiusesene

## AUTHOR CONTRIBUTIONS

VMK, BT, and IE conceived the article. IE is the guarantor. VMK, BT, AD, MNYY, YOM, NEK, FS wrote the manuscript. VMK, BT, AD, MNYY, YOM, NEK, FS, MNY, EF, TKF, YMP, OO, NR, YO, N, IE provided a critical appraisal of the manuscript. All the authors critically revised and approved the final manuscript.

## DATA AVAILABILITY STATEMENT

### FUNDING

None.

## COMPETING INTERESTS

None.

## Notes

### Competing Interest Statement

The authors have declared no competing interest.

